# Characteristics of Cerebrospinal Fluid in Autism Spectrum Disorder - A Systematic Review

**DOI:** 10.1101/2024.02.13.24302752

**Authors:** Vandana Srivastava, Christian O’Reilly

**Affiliations:** AI Institute, University of South Carolina, 5th floor, 1112 Greene St., Columbia, South Carolina 29201, USA; Department of Computer Science and Engineering, University of South Carolina, 550 Assembly Street, Columbia, South Carolina 29201, USA; Carolina Autism and Neurodevelopment Research Center, University of South Carolina, 1800 Gervais Street, Columbia, South Carolina 29201, USA; Institute for Mind and Brain, University of South Carolina, 1800 Gervais Street, Columbia, South Carolina 29201, USA

**Keywords:** autism spectrum disorder, cerebrospinal fluid, immune system, folate deficiency, neurotransmitters

## Abstract

Autism Spectrum Disorder (ASD) is a range of neurodevelopmental conditions characterized by impaired social interaction, learning, and restricted or repetitive behaviors. The underlying causes of ASD are still debated, but researchers have found many physiological traits like gut problems and impaired immune system to help understand the etiology of ASD. Cerebrospinal fluid (CSF) plays a critical role in maintaining the homeostasis of the neuronal environment and has, therefore, been analyzed in multiple conditions impacting the central nervous system. The study of CSF is crucial to understanding neurological disorders as its composition changes with the disorders, and these changes may indicate various disorder-related physiological mechanisms. For this systematic review, we searched PubMed, Scopus, and Web of Science for studies published between 1977 and 2025 and selected 49 studies after manual screening. We took stock of the evidence supporting the hypothesis that ASD alters the properties and composition of CSF. We systematically report on the different attributes of CSF in the ASD population that could be potential biomarkers and assist in understanding the origins and progression of ASD.

We found that in CSF, immune markers, proteins, extra-axial CSF, folate, oxytocin, and vasopressin showed changes in ASD compared to the neurotypicals. We observed gaps in the literature due to variations in age and sample size and noted biases related to sex (i.e., samples are predominantly including male participants) and age (i.e., a handful of studies were conducted on adults). Our review highlights the need for more research on CSF in ASD to improve our understanding of this disorder and identify CSF biomarkers.

## 1. Introduction

Autism Spectrum Disorder (ASD) is a complex neurodevelopmental disorder that results in an array of challenges associated with social interactions, communication, learning, and repetitive behaviors. Individuals with ASD may present varying etiologies, phenotypical profiles, and degrees of impairment. Hence, this condition is referred to as a “spectrum” disorder. According to the Centers for Disease Control and Prevention, the prevalence of ASD among children aged 8 years in the USA was 1 in 36 (2.8% overall; 4.3% of boys and 1.1% of girls) in 2020 (CDC, 2022). The factors contributing to the etiology of ASD are diverse and not fully ascertained. Still, many studies have been conducted to determine the anatomical and physiological characteristics specific to the ASD population. Analyzing the properties of the cerebrospinal fluid (CSF) in that population is one avenue that has been investigated to gain a deeper understanding of ASD.

The CSF is a clear, colorless fluid present in the region surrounding the brain and spinal cord of vertebrates (Adigun, Al-Dhahir, 2022). It serves multiple purposes, including keeping the brain floating, cushioning it from jolts and preventing associated trauma, helping distribute various substances between brain cells, and carrying away the waste produced from neural activity. CSF is found in the brain ventricles and the cranial and spinal subarachnoid space. It is produced mostly by the neuroepithelial lining (called *ependyma*) of the brain ventricular system (Jiménez et al., 2014) and the central canal of the spinal cord. These special ependymal cells exist in the choroid plexus, an organ that facilitates the entry of immune cells into the central nervous system (CNS) and monitors the synthesis, formation, and flow of CSF (Lun et al., 2015). The brain interstitial fluid and capillaries also secrete a small amount of CSF (Sakka et al., 2011).

The CSF is regenerated about 4-5 times per day. It is similar in composition to blood plasma except that it contains negligible proteins and has higher concentrations of Na^+^, Cl^−^, and Mg^+^, and lower concentrations of K^+^ and Ca^2+^ (Sakka et al., 2011). Monoamines like dopamine, serotonin, melatonin, and neuropeptides like atrial natriuretic peptide (ANP) play an important role in CSF regulation (Faraci et al., 1990). Along with monoamines and neuropeptides, arginine vasopressin (AVP) receptors are present on the surface of the choroidal epithelium. CSF secretion is found to be decreased by ANP and AVP (Faraci et al., 1990).

CSF is drawn invasively from the spinal cord, commonly through lumbar puncture, and carries some post-procedure risks like headache, local skin infection, and nausea (Czarniak et al., 2023). However, due to its proximity to CNS, properties of the CSF like the white blood counts, protein levels, and serum-glucose ratio can help differentiate CNS infections caused by distinct pathogens (Gomes, 2022), making it an important medical diagnostic tool. The circulation of CSF can also be determined non-invasively using Magnetic Resonance Imaging (MRI). The study and interpretation of CSF are critical to understanding neurological disorders as their composition, quantity, and flow change with the disorders (Hrishi & Sethuraman, 2019).

Further, since the level of several CSF makers like the homovanillic acid, 5-methyl-tetra-hydrofolate, and 5-hydroxyindoleacetic acid change very little under common storage and freeze/thaw conditions (Willemse et al., 2019), CSF can be analyzed reliably for biomarker studies.

With this review, we aim to synthesize the knowledge gathered from previous studies to clarify if and how ASD is associated with changes in the properties of the CSF. More specifically, we aim to thoroughly assess the evidence supporting the hypothesis that the composition and properties of the CSF are altered in ASD compared to neurotypical individuals. To the best of our knowledge, this is the first time a systematic review has been done to study the changes in different properties of CSF in ASD.

## 2. Methods

The search strategy followed the Preferred Reporting Items for Systematic Reviews and Meta-Analyses (PRISMA) guidelines (Figure 1). PubMed and Web of Science were queried with the research string *(“cerebrospinal fluid” or csf) and (“Autism” or “ASD”) not diet not schizo* not antisiphon not hydrocep\** to filter out the studies related to schizophrenia, hydrocephalus, and antisiphon devices. The Scopus database was queried with an equivalent research string. The results were limited to papers published between 1977 and March 2025. Papers were screened for relevance to this review using the following criteria:

1. CSF was analyzed in the context of ASD.
2. The paper was not a neuroimaging study, except when the focus was to characterize extra-axial CSF. For example, papers mentioning CSF but using magnetic resonance spectroscopy to assess in ASD the concentration of some molecules (e.g., GABA) not in the CSF were rejected.
3. The paper was not a review, a meta-analysis, or a case report.
4. The study was conducted on humans. Animal models were not included to ensure a sufficiently focused scope and to avoid potential issues associated with a lack of translatability of results between animal models and humans. Although not part of the material officially reviewed and not included in summary tables, we nevertheless mention relevant observations from animal studies in our discussion of the reviewed material.
5. The criteria used to identify the ASD group were either one or a combination of the following:

- Diagnostic and Statistical Manual of Mental Disorders-IV (DSM),
- Autism Diagnostic Interview-Revised (ADI-R),
- Autism Diagnostic Observation Schedule (ADOS),
- Clinical diagnosis by experts.

**Figure 1:**
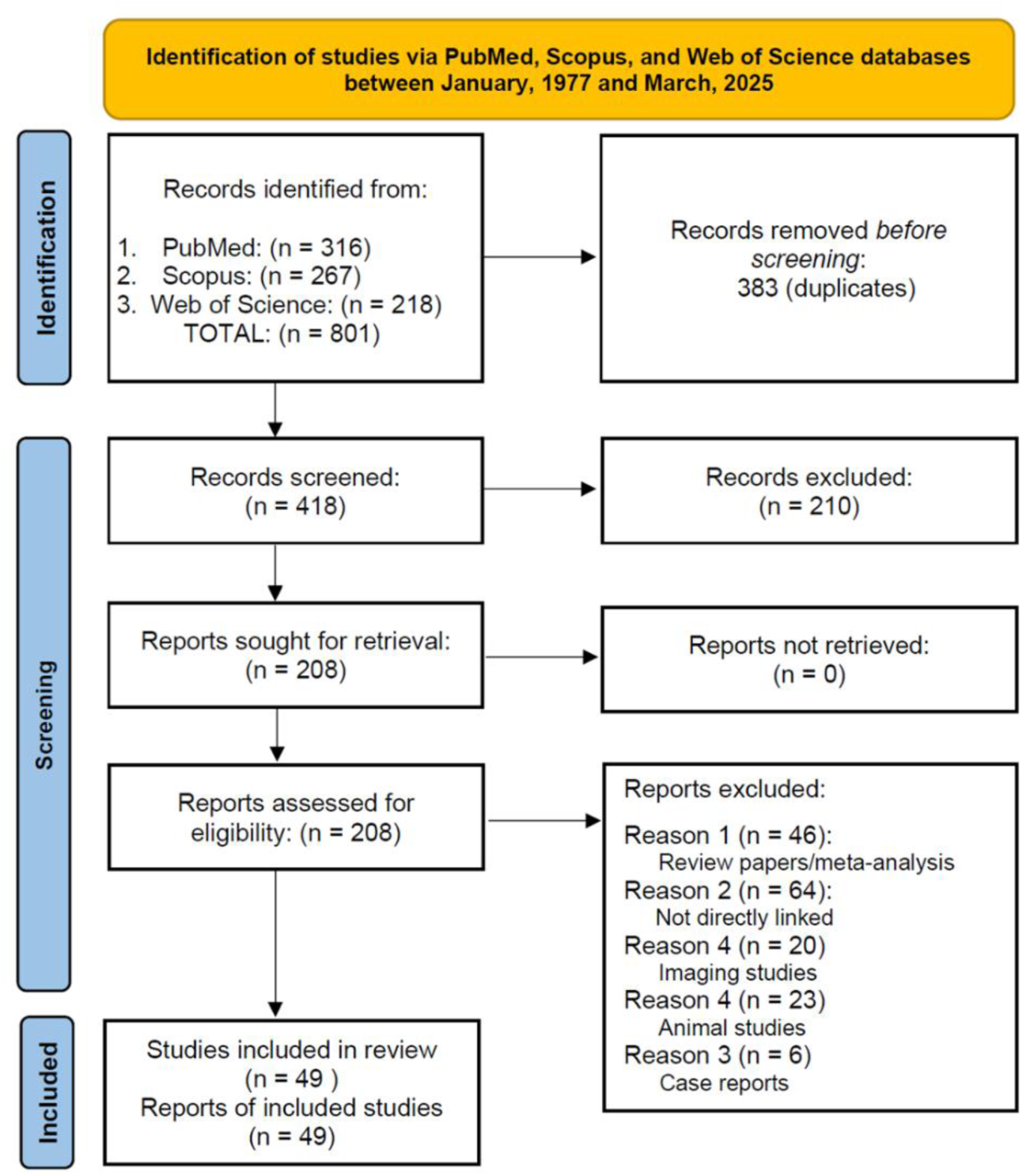
PRISMA flowchart describing the paper selection process.

The papers were manually reviewed and screened by the first author (VS). Problematic cases were discussed or independently reviewed by the senior author (COR). Out of a total of 801 papers, we discarded 383 duplicate papers and screened 418 papers. We shortlisted 49 papers that fulfilled the selection criteria and were considered for this review. The following data were collected from the studies and are systematically reported in Tables 1-6:

1. Author and year,
2. Age of both study group (ASD participants) and control group (non-ASD or neurotypical),
3. Size of the study group (count of ASD participants whose CSF is analyzed) and sex distribution (number of males and females) in the study group,
4. Size of the control group (count of non-ASD or neurotypicals whose CSF is analyzed) and sex distribution (number of males and females) in the control group,
5. Observations (CSF characteristics in the study group as compared to the control group).

We manually collected the data from each study from the “abstract” and “results” sections and organized it in tabular form. We used dashes (−) to denote missing values in the age/size column. To reduce the chances of bias, we included studies with null results.

## 3. Results

A total of 49 papers were studied to find the characteristics of CSF in ASD. We organized the papers into six categories based on their observed CSF properties. The categories are (percentage of papers in parenthesis):

1. Immune markers (13.7%)
2. Extra-axial CSF (23.6%)
3. Folate deficiency (11.8%)
4. Protein/amino acids (17.6%)
5. Monoamine neurotransmitters (serotonin, dopamine) (17.6%)
6. Others (oxytocin, arginine vasopressin, beta-endorphin, metabolites, and gangliosides) (15.7%)

**Two studies (Ramaekers et al., 2020) and (Young et al., 1977) were mentioned in two different tables (“monoamine neurotransmitters”, “folate deficiency”) and (“immune markers”, “folate deficiency”), respectively.**

The body of literature reviewed is biased concerning age and sex. Most of the studies included either only males or a large proportion of males in their research. Also, most human studies were conducted with infants or children, with very few studies on adults. We have reported the CSF results, as mentioned in the papers. As this review is not a meta-analysis and did not attempt to statistically aggregate reported results, we did not normalize or correct the CSF results for variation due to age or sex.

### 3.1 Presence of neuroinflammation markers in CSF

Many comorbidities of ASD indicate an altered immune system, including allergies (Xu et al., 2018), gastrointestinal issues (Saurman et al., 2020), and autoimmune diseases (Hughes et al., 2018). These comorbid conditions add to the ASD burden, often resulting in a poorer quality of life. The molecular properties of CSF change during inflammation and disease of the CNS (Świderek-Matysiak et al., 2023). In a neuro-inflammatory study including 75 participants, the level of 26 (in bold in the following list) out of 36 cytokines (**CCL1**, CCL2, **CCL3**, **CCL7-8**, **CCL11, CCL13, CCL19**-**20**, **CCL22-23**, CCL24, **CCL25**, CCL26-27, **CXCL1**-**2,** CXCL5, **CXCL6**, **CXCL8**-**9**, **CXCL11–13**, CXCL16, **CX3CL1**, **IL-2**, IL4, **IL-6, IL-10**, **IL-16**, GM-CSF, **IFN-γ**, MIF, **TNFα**, and MIB1β) was elevated in the CSF of patients with neuro-inflammatory diseases, whereas only three cytokines (CCL3, CXCL8, and IL6) had elevated levels in the serum (Lepennetier et al., 2019). CSF cytokine concentrations were also positively correlated with CSF immune cell counts (CD4 and CD8 T cells, B cells, plasmablasts, monocytes, and natural killer (NK) cells) (Lepennetier et al., 2019). Inflammatory molecules have been shown to impact neurodevelopment, and early-life inflammation has been linked with neurodevelopmental disorders like ASD, cerebral palsy, epilepsy, and schizophrenia (Jiang et al., 2018). Similarly in ASD, a significant increase in proinflammatory cytokines and growth factors was found, especially the chemokine MCP-1 (12-fold increase) (Vargas et al., 2005). Altered cytokines (IFN-γ, MCP-1, IL-8; chemokine subgroup) were also reported (Runge et al., 2023) in adults with ASD, but this study found decreased levels of MCP-1, IFN-γ and increased levels of IL-8. MCP-1 is a chemokine that is associated with innate immune reactions and is vital for monocyte and T-cell activation in regions of tissue injury (Vargas et al., 2005).

*Growth factors* play a regulatory role in the immune and vascular systems (Pardo et al., 2017). In a longitudinal study, a significant difference was found in CSF growth factors EGF and sCD40L (Pardo et al., 2017) in children (2 to 8 years of age) with ASD compared to controls. EGF is important for the growth, proliferation, and differentiation of numerous cell types and is involved in several pathways of neuronal function, whereas sCD40L modulates the function of B cells (Pardo et al., 2017). The CSF profiles of cytokines, chemokines, and growth factors did not change significantly in the follow-up performed one to three years later (Pardo et al., 2017). The same study found decreased levels of quinolinic acid, which indicates the inefficiency of the kynurenine pathway to form quinolinic acid from tryptophan (Zimmerman et al., 2005). Decreased neopterin in CSF with increased biopterin suggested neuroinflammation in the Autism group (Zimmerman et al., 2005).

Tumor necrosis factor-alpha (TNF-α) is an inflammatory cytokine produced by macrophages/monocytes during severe inflammation (Idriss & Naismith, 2000), and its elevation is associated with several autoimmune diseases like rheumatoid arthritis, Crohn’s disease, and psoriasis (Parameswaran & Patial, 2010). The presence of inflammation and immune system over-activation in ASD is supported by reports of elevated levels of TNF-α in the CSF of an all-male cohort (Chez et al., 2007). Another study found elevated levels of TNF-α in the serum but not the CSF of a group of participants with ASD (Zimmerman et al., 2005). However, this study reported no significant correlation between the level of TNF-α in serum and CSF, and the use of small sample sizes (n=12) may be responsible for non-conclusive tests. The presence of neuroinflammation is further supported by a recent study that found elevated levels of TNF-α and interleukins in ASD (Than et al., 2023).

Despite immune dysregulation in ASD, initial studies found CSF immunoglobulin (antibodies) levels in the normal range in the ASD cohort when compared with previous studies (Young et al., 1977). Table 1 summarizes the studies related to CSF immune markers in ASD.

**Table 1:** CSF immune markers in ASD.

|  | **Age (study group/control group)** | **Study group (ASD participants)** | **Control group (Non-ASD participants)** | **Observations (in the study group)** |
| --- | --- | --- | --- | --- |
| Than et al. 2023 | 3 - 11 years /<br>1 - 18 years | 26 (-) | 8 (-) | Significantly increased TNF-α, IL-4, IL-21 levels in CSF |
| Runge et al 2023 | 29 - 50 years /<br>- | 24 (16 males, 8 females) | Compared with previous study | Significantly decreased IFN-γ-induced protein-10 and MCP-1 levels, and significantly higher IL-8 levels were detected in ASD |
| Pardo et al. 2017 | 2 - 8 years / - | 67 (55 males, 12 females) | 54 (41 males, 13 females) | Significant differences in the growth factors in the CSF of the ASD group with respect to controls; profiles of cytokines, chemokines, and growth factors did not change significantly in ASD in the follow-up years |
| Chez et al. 2007 | 2.5 - 9.7 years / - | 10 (all males) | Compared with previous studies | CSF TNF-α was significantly increased in comparison to serum-TNF-α |
| Zimmerman et al. 2005 | 2.7 - 10 years /<br>2 - 14 years | 12 (10 males, 2 females) | 15 (6 males, 9 females) | Elevated levels of TNF-α in the serum but not the CSF of the ASD group; quinolinic acid and neopterin were decreased, and biopterin was elevated in ASD group |
| Vargas et al. 2005 | 3 - 10 years /<br>12 - 45 years | 6 (4 males, 2 females) | 9 (3 males, 6 females) | Significantly increased MCP-1(12-fold increase); no differences in expression of TARC or TGF-β1; increased cytokines (IL-6, IFN-γ, IL-8, MIP1β, NAP-2, IFN-γ inducing protein-10) and angiogenin; increased growth factors (MIF, VEGF, LIF, osteoprotegerin, HGF, PARC, FGF-4, FGF-9, IGFBP3, and IGFBP4) |
| Young et al. 1977 | 3.8 - 9.1 years / - | 15 (11 males, 4 females) | Compared with previous studies | CSF immunoglobulin levels were within normal limits |

Abbreviations- BDNF: Brain-derived neurotrophic factor, FGF: Fibroblast growth factor, HGF: Hepatic growth factor, IFN-γ : Interferon-gamma, IGFBP: Insulin-like growth factor binding protein, IL: Interleukin, LIF: Leukemia inhibitory factor, MCP-1: Monocyte chemoattractant protein-1, MIP1β : Macrophage inflammatory protein-1 $\beta$ , MIF: Mesoderm inducing factor, NAP-2: Neutrophil activating peptide-2, TARC: Thymus and activation regulated chemokine, TGF- $\beta$ 1: Tumor growth factor- $\beta$ 1, TNF: Tumor necrosis factor, VEGF: Vascular endothelial growth factor

Reviewed immune studies analyzed CSF for immune markers using luminex/bead multiplexing immunoassay (BMIA) (43%), sandwich enzyme immunoassay (SEIA)/Enzyme-linked immunosorbent assay (ELISA) (43%), or single-radial immunodiffusion (SRID) (14%).

### 3.2 Extra axial CSF in infants later diagnosed with ASD

The presence of increased CSF volume in the subarachnoid space surrounding the cortical surface is a brain abnormality referred to as extra-axial CSF (EA CSF). Having a large volume of CSF forces the ventricles to widen, causing pressure on brain tissues. This can eventually lead to brain damage. Many studies have confirmed that children later diagnosed with ASD had EA CSF from 6 to 24 months compared to neurotypicals (Hallahan et al., 2009; McAlonan et al., 2005; Shen, 2018; Shen et al., 2018; Shen & Piven, 2017). Excess CSF has been associated with an enlargement of the head circumference in ASD (Denier et al., 2022; Shen et al., 2013, 2018). EA CSF has also been linked with enlargement of the perivascular spaces (fluid-filled channels that surround blood vessels in the brain) from 6 to 24 months in ASD, leading to sleep problems later in ages between 7 and 12 years (Garic et al., 2023). The amount of EA CSF at 6 months has also been claimed to be predictive of later ASD symptom severity (Shen et al., 2013). CSF volume was reported to stabilize before 4 years of age (Peterson et al., 2021a). No significant difference in CSF volume was found for adults in one study (Creasey et al., 1986), although a different study reported subarachnoid CSF/meningeal CSF volume to be significantly different in ASD compared to controls in a male-only cohort of mean age 15 years (Tate et al., 2007). Finally, a study involving infants at elevated likelihood for ASD given the presence of an older sibling with a diagnosis of ASD found the brain size (but not EA CSF) of the younger siblings to be correlated with the scores on the Social Communication Questionaries of the older sibling in the groups where the younger sibling was diagnosed with ASD at 24 months (Girault et al., 2022).

The polygenic risk score (PRS) is a metric that is used to estimate an individual’s genetic predisposition to a trait or disease, calculated using a genotype profile and relevant genome-wide association study (GWAS) data (Choi et al., 2020). An MRI study of ∼31,000 adults, including ASD group and controls in the age group 40 – 70 years, reported the PRS scores to be correlated with increased CSF volume (Mohammad et al., 2024). This recent study affirms that the CSF pathway is altered in ASD, and it may have some genetic connection also.

The results for the relationship between EA CSF and ASD are summarized in Table 2.

**Table 2:** Extra-axial CSF in ASD.

|  | **Age (study group/control group)** | **Study group (ASD participants)** | **Control group (Non-ASD participants)** | **Observations (in the study group)** |
| --- | --- | --- | --- | --- |
| Mohammad, S. et al 2024 | 40 - 70 years / 40 - 70 years | 18,381* | 27,969* | CSF volumes were significantly positively correlated with the PRS |
| Garic et al. 2023 | 6.5, 12.7, 24.7 months / 6.7, 12.6, 24.7 months | 47 (ELA; ASD+; 40 males, 7 females)<br>180 (ELA; ASD-; 102 males, 78 females) | 84 (TLA; ASD-; 55 males, 29 females) | Across all groups, enlarged PVS at 24 months was associated with greater EA CSF volume from ages 6 to 24 months and more frequent night-wakings at school age. |
| Girault et al. 2022 | 6 months | 42 ELA; ASD+ | 149 ELA; ASD- | no significant association between proband autistic traits (SCQ score) and sibling EA CSF at 6, 12, or 24 months (for both the ASD+ and ASD- groups) |
|  | 12 months | 39 ELA; ASD+ | 182 ELA; ASD- |  |
|  | 24 months | 42 ELA; ASD+ | 159 ELA; ASD- |  |
| Denier et al. 2022 | 19.9 – 37.3 years / 20.7 - 34.5 years | 120 (all males) | 136 (all males) | Increased head circumference due to EA CSF |
| Peterson et al. 2021 | 3 - 42 years / - | 97 (all males) | 92 (all males) | No group differences in EA CSF volume |
| Shen et al. 2018 | 2 - 4 years/ - | 159 (132 males, 27 females) | 77 (49 males, 28 females) | ASD group had an average of 15.1% more EA CSF than controls; enlarged head circumference in the ASD group |
| Shen et al. 2017 | 6 - 24 months/ - | 221 ELA (47 ASD+) | 122 TLA | Elevated levels of EA CSF in the ELA group than TLA; 18% more EA CSF at 6 months in ELA ASD+ than in the ELA group |
| Shen et al. 2013 | 6 - 9 months/ - | 33 ELA (22 males, 11 females) (10 ASD+) | 22 TLA (15 males, 7 females) | Significantly elevated EA CSF in ELA infants at 6-9 months and continued until 1.5 - 2 years; the amount of EA CSF at 6 months was predictive of the severity of ASD symptoms; enlarged brain at an early age in ELA. |
|  | 1 - 1.2 years/ - | 27 ELA | 16 TLA |  |
|  | 1.5 - 2 years/ - | 26 ELA | 16 TLA |  |
| Hallahan et al. 2009 | > 18 years / Mean age: 32 years | 114 (96 males, 18 females) | 60 (53 males, 7 females) | Significantly larger volume of peripheral CSF in ASD |
| Tate et al. 2007 | Mean age 14.7 years / 13.6 years | 34 (all males) | 26 (all males) | Significant group differences in the relationship between subarachnoid CSF/meningeal volume |
| McAlonan et al. 2005 | 10.2 - 13.8 years / 9.8 - 12.2 years | 17 (16 males, 1 female) | 17 (16 males, 1 female) | Increased total CSF volume in ASD |
| Creasey et al. 1986 | 18 - 39 years / 21 - 37 years | 12 (all males) | 16 (all males) | No significant group differences in the volume of CSF |
| Abbreviations- ASD+: later diagnosed with ASD, ASD-: later diagnosed without ASD, EA CSF: Extra axial cerebrospinal fluid, ELA: Elevated likelihood for ASD, PRS: Polygenic risk score, PVS: Perivascular spaces, TLA: Typical likelihood for ASD<br>* Number of study participants (ASD + Controls) = ~31, 000 with 52% males and 48% females |  |  |  |  |

### 3.3 Low folate level in the CSF of people with ASD

Folate is a vitamin (B9) essential for brain health. It supports the creation of DNA and RNA, the formation of neurotransmitters, and the development of the nervous system during pregnancy (Balashova et al., 2018; Gordon, 2009). The predominant form of folate in cerebrospinal fluid is 5-methyl-tetra-hydrofolate (5-MTHF). Cerebral folate deficiency is caused by a malfunction in the folate receptor autoantibodies (FRA), a protein that binds to folate (Gordon, 2009). FRA is created in the choroid plexus localized within the border of the cerebral ventricles and moves into the CSF (also released by the choroid plexus) to be transported to the brain. Cerebral folate deficiency can be caused by the presence of FRA, which interferes with the function of that receptor.

The earliest study (Lowe et al., 1981) conducted on autistic and non-autistic neuropsychiatric patients found the CSF folate levels to be within the normal range in both groups. The subset of patients in the autistic group who were given oral folic acid supplements did not show clinical improvements (Lowe et al., 1981). Similarly, a more recent longitudinal study (two time points separated by 1-3 years) exploring the association between cerebral folate deficiency and autism found no significant correlation between CSF 5-MTHF levels and autistic features (Shoffner et al., 2016). However, a few studies found low CSF 5-MTHF in ASD (Frye et al., 2013; Ramaekers et al., 2007) and proposed oral *d,l-leucovorin (folinic acid)* (Frye et al., 2013; Ramaekers et al., 2007) to alleviate ASD-associated symptoms. In some cases (Frye et al., 2013, 2020; Ramaekers et al., 2007), oral folic acid improved verbal communication, motor skills, and CSF 5-MTHF levels in the ASD group. Frye et al. (2013) reported the presence of FRA in 75.3% (70/93) of the ASD group. A recent study found the CSF to have lower folate levels in only 21% (8/38) of ASD participants when compared to controls (Ramaekers et al., 2020). Table 3 summarizes the CSF folate deficiency papers in ASD.

**Table 3:** CSF folate levels in ASD.

|  | **Age (study group/control group)** | **Study group (ASD participants)** | **Control group (Non-ASD participants)** | **Observations (in the study group)** |
| --- | --- | --- | --- | --- |
| Ramaekers et al., 2020 | 3.6 - 18 years / 4.4 - 16 years | 38 (31 males, 7 females) | 24 (12 males, 12 females) | Lower 5-MTHF in 21% of participants with ASD; FRAA present in 68% of ASD group |
| Shoffner et al., 2016 | 2 - 6 years / - | Visit 1: 67 (-) | 140 (-) | CSF 5-MTHF levels vary significantly over time in an unpredictable way; no significant relationship to clinical features of autism |
|  | 3.5 - 9.5 years / 2 - 10 years | Visit 2 (1-3 years later): 31 (-) |  |  |
| Frye et al., 2013 | 2.9 - 17.4 years / - | 93 (84 males, 9 females) | 44 treated with leucovorin<br>26 untreated | High prevalence (75.3%) of FRA; low CSF 5-MTHF; improvement in treated vs. untreated children on verbal communication, receptive and expressive language, attention |
| Ramaekers et al., 2007 | 2.8 - 12.3 years / 3.3 - 11.4 years | 25 (18 males, 7 females) | 100 (56 males, 44 females) | CSF 5-MTHF was low in 23 (92%) participants with ASD; oral folic acid supplements led to normal CSF 5-MTHF levels |
| Lowe et al., 1981 | 4 - 22 years / 2 - 32 years | 16 (-) | 19 (-) | CSF folate levels were within the normal range in both groups |
| Young et al. 1977 | 3.8 - 9.1 years / - | 15 (11 males, 4 females) | Compared with previous studies | CSF folate levels were within normal limits |
| Abbreviations- FRA: Folate receptor autoantibodies, MTHF: methyl-tetra-hydrofolate |  |  |  |  |

Reviewed folate studies analyzed CSF for folate levels using high-performance liquid chromatography (HPLC) / Reverse-phase High-performance liquid chromatography (RP-HPLC) (33%), aseptic addition method (AA) (17%), ELISA (33%), and SRID (17%).

### 3.4 Changes in the CSF protein levels associated with ASD

Amino acids are molecules that combine to form proteins. These proteins serve many cellular functions. A large proportion of proteins in normal CSF is derived from blood, e.g., albumin which constitutes 35 - 80% of total protein in CSF. About 20% of the proteins in CSF are produced in the brain by neurons, glial cells, and leptomeningeal cells (Reiber, 2003). Changes in the brain-derived CSF protein concentration may indicate CNS disorders, impaired blood-brain barrier, or disruption in CSF flow (Reiber, 1994, 2003; Schilde et al., 2018; Wichmann et al., 2021). The CSF analysis in ASD found altered levels of albumin and several other proteins. In adults with ASD, the overall protein concentration and albumin quotient (ratio of CSF and serum albumin (Andrews et al., 1994)) were found to be increased (Runge et al., 2020). An elevated concentration of ethanolamine (Perry, 1978) suggested CNS abnormality in ASD. Astrocytes are glial cells that ensure the defense and support of CNS during development, across adulthood, and in aging. Glial fibrillary acidic protein (GFAP) and S-100 are two of the many proteins expressed in astrocytes that provide strength to glial cells and maintain the blood-brain barrier (Kuroda, 1983). In response to brain injury or other neuro-damaging conditions, astrocytes trigger processes (reactive astrogliosis) that change the level of GFAP (Verkhratsky & Nedergaard, 2018; Yang & Wang, 2015). The CSF GFAP levels were found to be higher in ASD compared to controls (Ahlsén et al., 1993; Rosengren et al., 1992). In contrast, no group differences were found for S-100 protein concentrations (Ahlsén et al., 1993; Rosengren et al., 1992).

To find the association between the CSF proteins and autistic traits in twins diagnosed with ASD and other neurodevelopmental disorders, a study measured 203 proteins in cerebrospinal fluid (n = 86, ASD = 19, neurotypical = 41). The autistic traits correlated significantly with four CSF proteins (Smedler et al., 2021):

1. C-C motif chemokine ligand 23 (CCL23) – a chemokine active on immune cells like T lymphocytes and monocytes (Karan, 2021).
2. Agouti-related protein (AGRP) – a protein synthesized in hypothalamic neurons and involved in energy metabolism and appetite.
3. Chitinase-3-like protein 1 (CHI3L1) – protein marker of inflammation.
4. Lipopolysaccharide-induced TNF-α factor (LITAF).

CCL23, AGRP, and CHI3L1 correlated negatively, and LITAF correlated positively with autistic traits. Within twin pairs, no CSF protein concentrations were significantly associated with autistic traits.

Proteins such as insulin-like growth factors, IGF-1, and IGF-2 are involved in the growth and development of the nervous system. In ASD, low CSF IGF-1 concentration was detected (Riikonen et al., 2006; Vanhala et al., 2001), but no difference was found in CSF IGF-2 levels (Riikonen et al., 2006). Nerve growth factor (NGF), a protein similar to insulin (Andres & Bradshaw, 1980), is vital for the development and maintenance of sympathetic, sensory, and forebrain cholinergic neurons (Aloe et al., 2015). A study evaluating the CSF NGF concentrations found no significant differences in ASD compared to controls (Riikonen & Vanhala, 1999).

In the CSF of three children with ASD, including two siblings, succinyladenosine and succinyl-aminoimidazole carboxamide riboside purines were found (Jaeken & Van den Berghe, 1984), indicating a deficiency in the adenylosuccinate enzyme in the brain of at least a subgroup of individuals with genetically defined ASD (Jurecka et al., 2015). This enzyme is involved in the synthesis of purines and adenosine monophosphate. Table 4 summarizes the results related to amino acids/protein concentration in the CSF of individuals with ASD.

**Table 4:** CSF protein levels in ASD.

|  | **Age (study group/control group)** | **Study group (ASD participants)** | **Control group (Non-ASD participants)** | **Observations (in the study group)** |
| --- | --- | --- | --- | --- |
| Smedler et al. 2021* | 8 - 23 years / 8 - 23 years | 19 (-) | 41 (-) | Twin study; in CSF across individuals, autistic traits correlated negatively with CCL23, AGRP, and CHI3L1, and positively with LITAF; within twin pairs, no CSF protein concentrations were significantly related with autistic traits |
| Runge et al. 2020 | 18 - 53 years / 18 - 61 years | 36 (23 males, 13 females) | 39 (6 males, 33 females) | Increased protein concentrations and albumin quotients in ASD |
| Riikonen et al. 2006 | 1.9 - 15.9 years / - | 25 (20 males, 5 females) | 16 (8 males, 8 females) | IGF-1 concentration was significantly lower in ASD; head circumferences correlated with IGF-1; no group differences in IGF-2 concentrations |
| Vanhala et al. 2001 | 1.9 - 6.5 years / 1.9 - 5.5 years | 11 (7 males, 4 females) | 11 (5 males, 6 females) | Levels of IGF-1 were significantly lower in ASD |
| Riikonen et al. 1999 | Mean age: 3.9 years / 4.3 years | 14 (9 males, 5 females) | 24 (12 males, 12 females) | No significant between-group differences in CSF NGF |
| Ahlsén et al. 1993 | 1.2 - 16 years / 1.3 - 18 years | 47 (32 males, 15 females) | 10 (7males, 3 females) | No group differences in CSF S-100 protein; CSF GFAP was ~3x higher in ASD |
| Rosengren et al. 1992 | 1.2 - 16 years / 1.3 - 29 years | 47 (32 males, 15 females) | 13 (8 male, 5 females) | Higher CSF GFAP levels in ASD; similar S-100 protein concentrations in both groups |
| Jaeken and Berghe 1984 | 1.6 - 3.7 years / - | 3 (2 males, 1 female) | 82 (-) | CSF concentration of succinyladenosine and succinylaminoimidazole carboxamide riboside were significantly increased in ASD |
| Perry et al. 1978** | 2 - 23 years / 2 - 15 years | 16 (-) | 23 (-) | The mean concentration of ethanolamine in CSF was significantly elevated in ASD |
| Abbreviations- AGRP: Agouti-related protein, BCAA: Branched-chain amino acids, CCL23: C-C motif chemokine ligand 23, GFAP: Glial fibrillary acidic protein, GST: Guggulsterone, IGF: Insulin-like growth factor, LITAF: Lipopolysaccharide-induced TNF-α factor, NGF: Nerve growth factor; PPA: Propionic acid<br>* ASD group (19/86); Control group (41/86); Males (55/86); Females (33/86)<br>** ASD group (28/34); Males (19/28): CSF analyzed in ASD group (16/19) |  |  |  |  |

Reviewed protein/amino-acid studies analyzed CSF for identifying protein properties using SEIA/ELISA (45%), fixed cell (FC) (11%), gas chromatography (GC) (11%), radioimmunoassay (RIA) (11%), MIA/BMIA (11%), or dual wavelength spectrophotometer (SP) (11%).

### 3.5 Monoamine neurotransmitter synthesis in ASD

Serotonin and the catecholamines dopamine, adrenaline, and noradrenaline are all important monoamine neurotransmitters. These compounds are involved in many CNS functions, including motor control, cognition, and emotion, as well as autonomic functions such as cardiovascular, respiratory, and gastrointestinal control (Pons, 2010). The synthesis of serotonin and dopamine leads to the formation of 5-hydroxyindolacetic acid (5-HIAA) and homovanillic acid (HVA), respectively (Lenchner JR, & Santos C., 2023). Changes in the levels of 5-HIAA and HVA are associated with aggressive, impulsive, and depressive behavior (Seo et al., 2008). In ASD, the CSF HVA was found to be increased (Gillberg et al., 1983; Gillberg & Svennerholm, 1987; Komori et al., 1995; Toda et al., 2006), indicating a disturbance in dopamine synthesis. However, no significant difference was reported for 5-HIAA (Gillberg et al., 1983; Gillberg & Svennerholm, 1987; Komori et al., 1995). A later study found no significant group difference in HVA and 5-HIAA levels (Narayan et al., 1993). Extending this study to a larger ASD cohort (Adamsen et al., 2014), low 5-HIAA levels were found in 56% (26/46) participants. A 2020 study also reported reduced 5-HIAA concentration compared to controls in 34% (13/38) of the ASD participants (Ramaekers et al., 2020).

Tryptamines, such as the serotonin and melatonin neurotransmitters, are derived from the essential amino acid, tryptophan. Tryptamine is a trace amine that activates amine-associated receptors in the brain of mammals and regulates the activity of dopaminergic, serotonergic, and glutamatergic systems (Gainetdinov et al., 2018). Indoleacetic acid, a tryptamine metabolite, was found in typical concentration in the CSF of people with ASD (Anderson et al., 1988), suggesting that the central metabolism of tryptamine is likely normal in ASD. However, differences in the tryptophan metabolism were reported in ASD (Boccuto et al., 2013; Kałużna-Czaplińska et al., 2017) particularly concerning the kynurenine pathway (Bryn et al., 2017; Carpita et al., 2023; Launay et al., 2023), but CSF analyses of metabolites generated by the kynurenine pathway are lacking.

Tetrahydrobiopterin (BH4) participates in the synthesis of monoamine neurotransmitters like dopamine, noradrenaline, and serotonin. This compound contributes to cellular metabolic pathways generating energy and protecting cells from inflammation (Eichwald et al., 2023). Studies have shown that ASD is related to the dysfunction of the cerebral dopaminergic and serotonergic systems (Nakamura et al., 2010). The CSF BH4 levels were found to be significantly reduced in ASD compared to controls (Tani et al. 1994). Low serotonin levels (5-HIAA) in CSF were unchanged when 6R-L-erythro-5,6,7,8-tetrahydrobiopterin (R-THBP) was used as an oral therapy (Komori et al., 1995), but half of the ASD group (7/14) showed improvement in autistic traits. Secretin, a digestive hormone, was found to promote the metabolism of serotonin and dopamine in the CNS and improved speech and sociability in 58% (7/12) of the participants with ASD and elevated CSF HVA levels (Toda et al., 2006). Table 5 summarizes the results related to monoamine neurotransmitters.

**Table 5:** CSF monoamine neurotransmitters in ASD.

|  | **Age (study group/control group)** | **Study group (ASD participants)** | **Control group (Non-ASD participants)** | **Observations (in the study group)** |
| --- | --- | --- | --- | --- |
| Ramaekers et al., 2020 | 3.6 - 18 years / 4.4 - 16 years | 38 (31 males, 7 females) | 24 (12 males, 12 females) | Lowered 5HIAA in 34% of ASD group; low 5HIAA to HVA ratio in 13% of ASD group |
| Adamsen et al. 2014 | - | 98 (-) | CSF not analyzed | Low 5HIAA in CSF in 26 (27%) ASD participants |
| Toda et al. 2006 | 4 - 16 years / 3 - 15 years | 12 (8 males, 4 females) | 17 (12 males, 5 females) | Increased HVA (9; 75%), 5-HIAA (7; 58%), R-BH4 levels (7; 58%) in the CSF of participants with ASD and improved autistic symptoms after secretin administration |
| Komori et al. 1995 | 2 - 9 years / 2.8 - 7 years | 14 (7 males, 7 females) | 18 (17 males, 1 female) | Increased level of mean HVA in ASD; no group differences in 5-HIAA; 7 (50%) participants with ASD showed improvements in autistic traits after R-THBP therapy |
| Tani et al. 1994 | 2.3 - 22.5 years / 0 - 12 years | 20 (15 males, 5 females) | 10 (7 males, 3 females) | NH<sub>2</sub> and R-BH4 levels in children with ASD were reduced to 66.1% and 41.5% with respect to controls |
| Narayan et al. 1993 | 2.9 - 8.5 years / 6.1 - 11.5 years | 17 (12 males, 5 females) | 15 (11 males, 4 females) | No group differences in the group mean for 5HIAA and HVA levels |
| Anderson et al. 1988 | 5.2 - 10.8 years / 3.7 - 9 years | 8 (7 males, 1 female) | 10 (all males) | No group differences in CSF levels of the tryptamine metabolite and IAA |
| Gillberg et al. 1987 | 1 - 16 years / Mean age: 8.3 years | 25 (20 males, 5 females) | 20 (15 males, 5 females) | Increased mean HVA concentration in ASD; no group differences in 5-HIAA |
| Gillberg et al. 1983 | 3 - 14 years / 3 - 14 years | 13 (10 males, 3 females) | 13 (10 males, 3 females) | Increased level of HVA in ASD; no group differences in 5-HIAA |

Abbreviations- 5HIAA: 5-hydroxyindolacetic acid, HVA: Homovanillic acid, IAA: Indoleacetic acid, NH<sub>2</sub>: 7,8-dihydroneopterin, R-BH<sub>4</sub>: 6R-5,6,7,8-tetrahydrobiopterin

Reviewed monoamine neurotransmitter studies analyzed CSF for the monoamine levels using HPLC/RP-HPLC (67%), mass fragmentographic (MF)(22%), or bicinchoninic Acid (BCA) protein assay (11%).

### 3.6 Changes in the levels of oxytocin, arginine vasopressin, beta-endorphin, metabolites, and gangliosides in the CSF of people with ASD

Oxytocin (OT) and arginine vasopressin (AVP) are nonapeptides that can cross the blood-brain barrier. These peptides are mainly synthesized by neurons of the paraventricular nucleus (PVN) and the supraoptic nucleus (SON) of the hypothalamus (Higashida, 2016; Jin et al., 2007). Studies have demonstrated that AVP/OT neurons extend processes that cross the walls of the third ventricle to release these neuropeptides directly into the CSF (Grinevich et al., 2016; Taub et al., 2021). Both these nonapeptides are found in the brains of males and females alike (Higashida et al., 2018; Neumann, 2008) and have an important role in neuronal function, social recognition, and social behavior in mammals, including humans (Higashida, 2016; Lukas & Neumann, 2013). Any change in the level of OT and AVP in CSF indicates impaired social behavior in mammals.

In OT/AVP studies on humans, the CSF AVP levels were significantly lower in an all-male ASD group (Parker et al., 2018) as compared to controls. The CSF AVP concentration was found to be decreased in the ASD group (24 males, 12 females), irrespective of sexes (Oztan et al., 2018). Intranasal vasopressin treatment improved autism-like symptoms in non-human primate (Parker, 2022). Research reported that infants who were later diagnosed with ASD had very low neonatal CSF AVP concentrations (Oztan et al., 2020). In an OT study with 0-3 months infants, no difference in neonatal CSF OT concentrations was found in infants later diagnosed with ASD compared to control. Altogether, the studies support the use of OT and AVP as CSF biomarkers in studying ASD or social symptoms in autism.

β-Endorphins are one of five groups of naturally occurring opioid peptides found in neurons of the hypothalamus and the pituitary gland. In stressful situations, the pituitary gland releases beta-endorphins into the CSF (Gifford & Mahler, 2012). Beta endorphins are associated with emotions like hunger, thrill, pain, and cognition (Pilozzi et al., 2020). Studies found no significant difference in beta-endorphin levels between the ASD and controls (Nagamitsu, 1993). In an earlier crossover study (Ross et al., 1987) to assess the effect of fenfluramine in infantile autism, beta-endorphin was reported to be elevated in ASD group.

Brain gangliosides play an important role in synaptic transmission, and increased synaptic activity leads to the release of more gangliosides (Lekman et al., 1995). Four major gangliosides (GM1, GD1a, GD1b, and GT1b) were found to be increased in the CSF of people with ASD (Lekman et al., 1995; Nordin et al., 1998).

Metabolomics is the study of metabolites, which are molecules associated with metabolism. This field has developed as a promising tool for understanding biochemical pathways in neurodevelopmental disorders (Yan et al., 2025). In the autistic regression cohort of 14, β-hydroxybutyrate was significantly decreased in the CSF, sphingolipids such as ceramides, hexosylceramides, and sulfatides were upregulated, and sphingomyelins were downregulated (Yan et al., 2025) in comparison to the 50 neurodevelopmental controls (without autistic regression). β-hydroxybutyrate is produced by the liver and is an essential carrier of energy from the liver to surrounding tissues when the glucose supply is low as compared to body energy needs (Newman & Verdin, 2017). People with ASD often have unregulated eating habits along with impaired gut health, which might lead to decreased butyrate production and lethargy (Yan et al., 2025). Similarly, an impaired sphingolipids pathway indicates autoimmunity in ASD, as sphingolipids are important for innate and adaptive immune system (Yan et al., 2025).

Reviewed CSF AVP studies used EIA/ELISA (38%) to analyze CSF. Beta-endorphin levels were measured by RIA method in 25% of the papers, modified microimmunoafinity procedure (MM) was used to measure brain gangliosides in 25% of the studies, and metabolites were measured using HPLC in 12% of the studies. Table 6 presents the results of ASD-related studies evaluating the concentration of nonapeptides, beta-endorphins, metabolites, and ganglioside levels in the CSF.

**Table 6:**
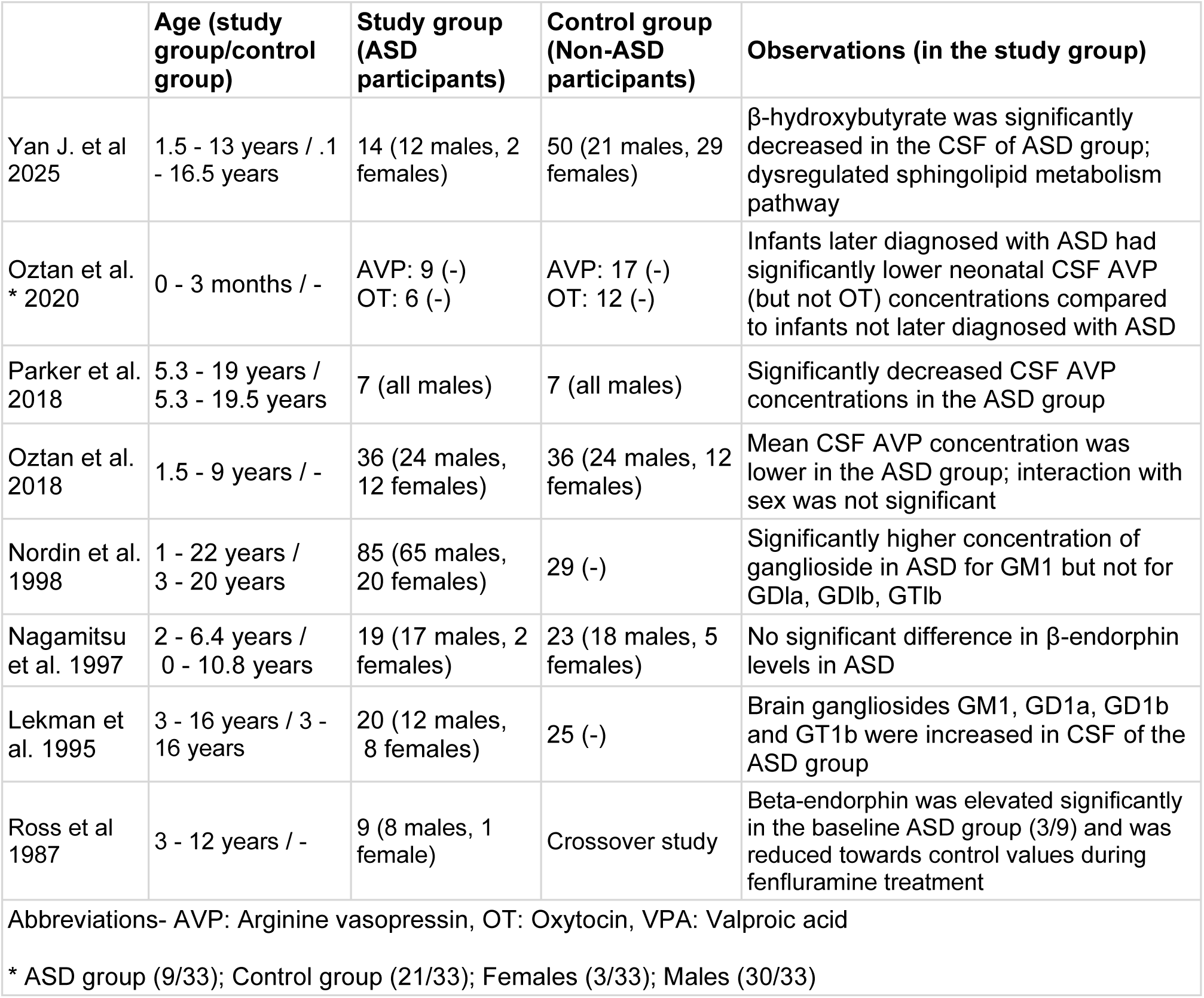
CSF concentration of OT, AVP, beta-endorphins, metabolites, and gangliosides in ASD.

## 4. Discussion

### 4.1 Summary of the findings

Our systematic review covers the research conducted to test whether the typical composition of CSF is altered in ASD. We found that several properties (e.g., immune markers, proteins, growth factors, folate level, and axial-CSF) get altered in ASD, whereas some properties (e.g., beta-endorphin, immunoglobulin, and indoleacetic acid) remain unchanged. Figure 2 gives a complete list of increased, decreased, and unchanged CSF properties according to the literature.

**Figure 2:**
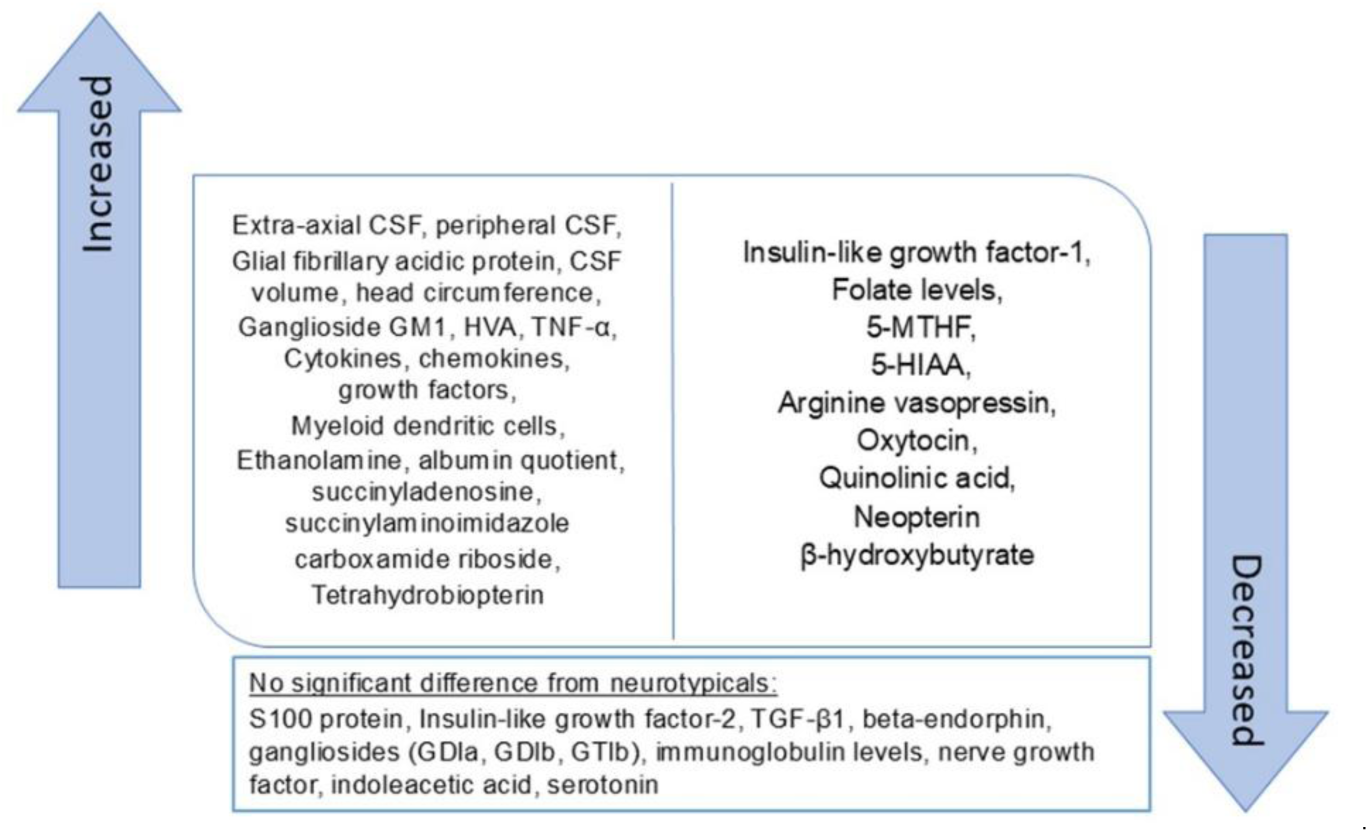
Summary of the CSF properties that are increased, decreased, or not significantly changed in ASD.

While reviewing the literature, we found that various methods were used to measure the chemical composition of the CSF. In the 37 CSF chemical analysis studies, the top three most frequently used methods were EIA/ELISA (12/37, 32%), followed by HPLC/RP-HPLC (8/37, 22%), and MIA/BMIA (4/37, 11%). According to the National Cancer Institute, an “assay” is defined as a laboratory test to find and measure the amount of a specific substance. Changes across time in assay techniques used in these studies are summarized in Figure 3. The HPLC methods are known to be very specific but costly and time-consuming in comparison to immunoassays (e.g., ELISA, RIA), which are cheaper and faster (Curwin et al., 2010). This may be the reason why most studies preferred immunoassay methods to determine the molecular composition of CSF.

**Figure 3:**
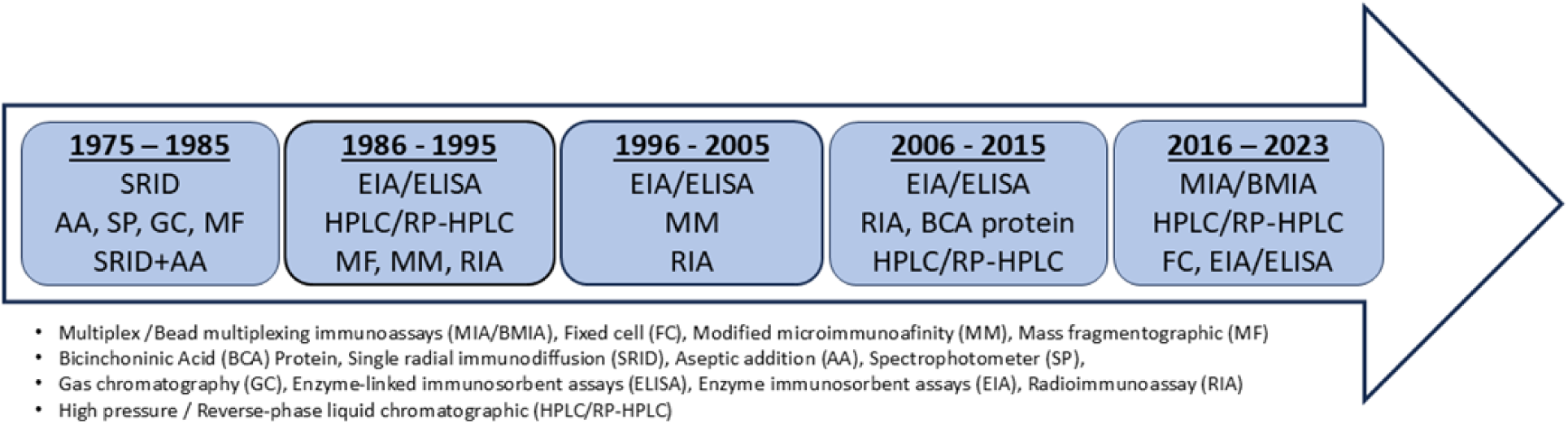
Assay methods used for analyzing chemical composition of the CSF

In 37 out of 49 studies reviewed, the CSF molecular (e.g., proteins, monoamines, folate) properties were tested. Figure 4 shows the percentage of molecules that changed significantly for each study, as defined by taking the ratio of the number of significantly changed molecule levels over the total number of molecules assayed. From Figure 4 (top and bottom), we found that six out of nine (67%) monoamine neurotransmitter studies were conducted before 1995, and 4/9 (44%) of them used the high-performance liquid chromatography (HPLC) method for analysis. 85% (6/7) of all the immune marker studies were carried out after 2000; 50% of them used multiplex/ bead multiplex immunoassays (MIA/BMIA), and 50% used the enzyme or enzyme-linked immunoabsorbent assays (EIA/ELISA) method. Also, 6/37 studies before 2000 were inconclusive (0%), but none were after 2000. This apparent increase in significant results may be due to the analysis methods used as well as the increase in sample sizes. The ASD sample size started increasing after 1995.

**Figure 4:**
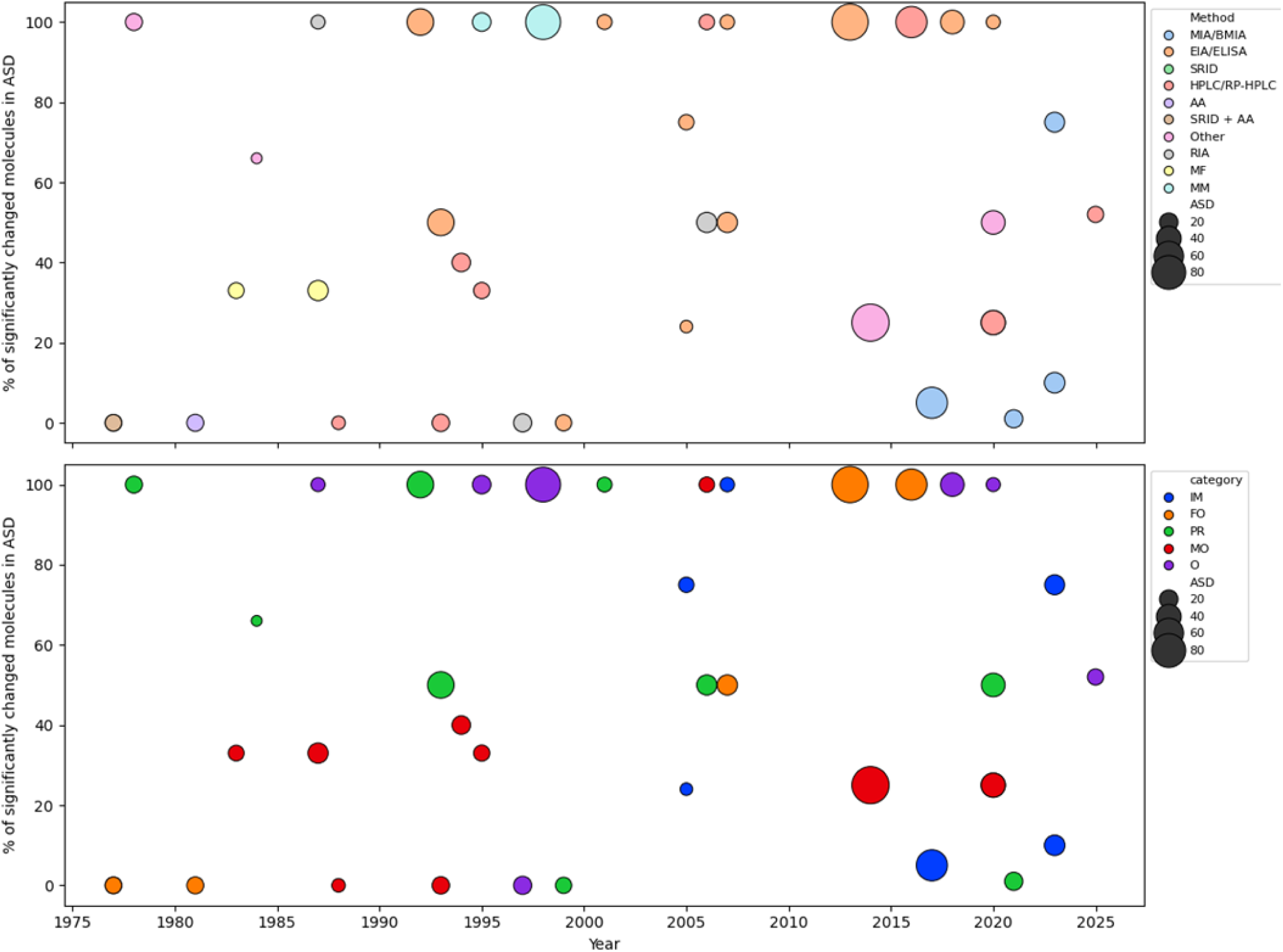
(top) Percentage of significantly changed molecules in ASD for 37 studies by year, sample size, and assay method. The “other” category in methods includes one study each of GC, SP, FC, and BCA. (bottom) Percentage of significantly changed molecules in ASD for 37 studies by year, sample size, and categories. The size of the circle represents the ASD group sample size, and the color represents the assaying methods (top) for analyzing CSF molecules and categories (bottom). The categories are IM: immune markers, FO: folate, MO: monoamine neurotransmitters, PR: protein/amino acid, and O: others

From Figure 4 (top), we did see that the percentage of significant molecules improved over the years, maybe due to better assaying methods. But when we tested the relation between PSM and (sample size, analysis method) and PSM with method, we did not find any statistically significant relation between PSM and (sample size + method) or PSM and method.

We also tested whether the percentage of significant molecules reported by studies was directly associated with their sample size for each of the five categories and found a 0.91 Pearson’s correlation that was statistically significant (p=0.012) for only the ‘folate’ group, even with this small sample of studies (n=6). We did not find any statistically significant relation between the percentage of significant molecules with ASD sample size for all the studies (n=37).

The systematic review provided clear evidence of increased cytokines (TNF-α, IFN-γ, IL-6, IL-8, MIP1β, NAP-2, IFN-γ), chemokines (MCP-1), and growth factors in the CSF of the ASD population. The elevated immune markers like cytokines, chemokines, and growth factors suggest an overactive innate immune system. Elevated CSF TNF-α but normal blood serum TNF-α indicates neuroinflammation. This unregulated inflammation can induce apoptosis or cellular death (Idriss & Naismith, 2000) and lead to inflammatory diseases (Van Loo & Bertrand, 2023). Also, the increased MCP-1 reported in children (Vargas et al., 2005) followed by decreased MCP-1 in adults (Runge et al., 2023) pointed out that maybe MCP-1 levels vary in ASD with age, but this hypothesis requires further corroboration.

ASD is also characterized by increased axial-CSF. Prior studies established that EA CSF increased between 6 months and 2 years of age in ASD. EA CSF at a young age indicates a disruption in CSF absorption in the first year after birth when CSF production is elevated (Murphy et al., 2020). The CSF increases till age 2 and then plateaus. This elevation results in increased head circumference, third ventricle, and total CSF volume in ASD compared to typically developing individuals. The increased EA CSF constitutes a promising biomarker for the early detection of ASD.

We found low folate levels reported in ASD in 50% (3/6) of the studies we reviewed (Frye et al., 2013; Ramaekers et al., 2020, 2007), with evidence of high FRA in one study (Frye et al., 2013). These results were contrary to the initial studies (Lowe et al., 1981; Young et al., 1977) where the folate levels were found to be within the normal range in both ASD and control groups. Folate deficiency has many side effects like psychomotor retardation, ataxia, and speech problems (Gordon, 2009), and can lead to depression (Gilbody et al., 2007).

Research on CSF proteins reported high GFAP in 22% (2/9) of the reviewed studies. Normally, CSF GFAP does not change much in comparison to plasma GFAP (Benedet et al., 2021). Hence, high CSF GFAP values can be a potential biomarker for ASD. We also found high protein/ethanolamine concentrations in 22% (2/9) and lower IGF-1 levels in 22% (2/7) of studies. No difference was observed for IGF-2, S-100, and NGF.

Four out of seven studies on CSF monoamine neurotransmitter (Gillberg et al., 1983; Gillberg & Svennerholm, 1987; Komori et al., 1995; Narayan et al., 1993) showed no significant difference in the mean serotonin level in CSF. In ASD, a lower level of serotonin was reported in two (Adamsen et al., 2014; Ramaekers et al., 2020) and higher levels in one (Toda et al., 2006) out of seven studies. Although a majority of studies did not report significant changes in serotonin, most recent studies (Adamsen et al., 2014; Ramaekers et al., 2020) showed low serotonin in CSF (34% and 26% of the participants with ASD, respectively), justifying more exploration. Furthermore, reduced BH4 (which synthesizes dopamine) and high HVA levels in 86% (6/7) of studies indicate a disturbance in dopamine synthesis in ASD.

Nonapeptide AVP was found to be decreased in infants (Oztan et al., 2020) and children/adults (Oztan et al., 2018; Parker et al., 2018). We found only three human studies on CSF related to OT and AVP. The effect of OT and AVP has mostly been studied on monkeys and rodent models. In the VPA-induced model of autism, pregnant rats were injected with valproic acid (VPA). The offspring of these VPA-exposed mothers show behavioral, immunological, and physiological changes similar to those described in the autistic population (Liu et al., 2018). CSF OT/AVP levels were altered in studies on monkeys and rat models of ASD. Low levels of OT in CSF (Dai et al., 2018; Gerasimenko et al., 2020) were found in adult VPA-induced rats and adult CD-157 knockout mice. The AVP studies established that CSF AVP concentrations correlate with social behaviors in monkeys (Oztan et al., 2021; Parker, 2022; Parker et al., 2018).

### 4.2 Research hypotheses derived from literature

The physiological characteristics of CSF in ASD led us to propose a few research hypotheses requiring further investigation. The following subsections discuss these hypotheses.

#### 4.2.1 Folate deficiency is linked to autistic characteristics

Folate + S-adenosylmethionine (SAMe) is crucial in regulating the production of THBP (or BH4), which is a co-factor in the synthesis of monoamine neurotransmitters (Bender et al., 2017). Thus, folate deficiency in ASD can be linked to low BH4, impaired monoamine synthesis, and high HVA. Research indicates that these deficiencies result in sleep disorders and disrupted psychomotor, social, and verbal skills (Galli et al., 2022; Sasa et al., 2003; Yoshimura et al., 2020), all traits associated with ASD. These observations explain the use of R-TBHP and folic acid as treatments to improve concentration, motor skills, and verbal communication in ASD. Figure 5 shows the relationship of ASD with folate deficiency.

**Figure 5:**
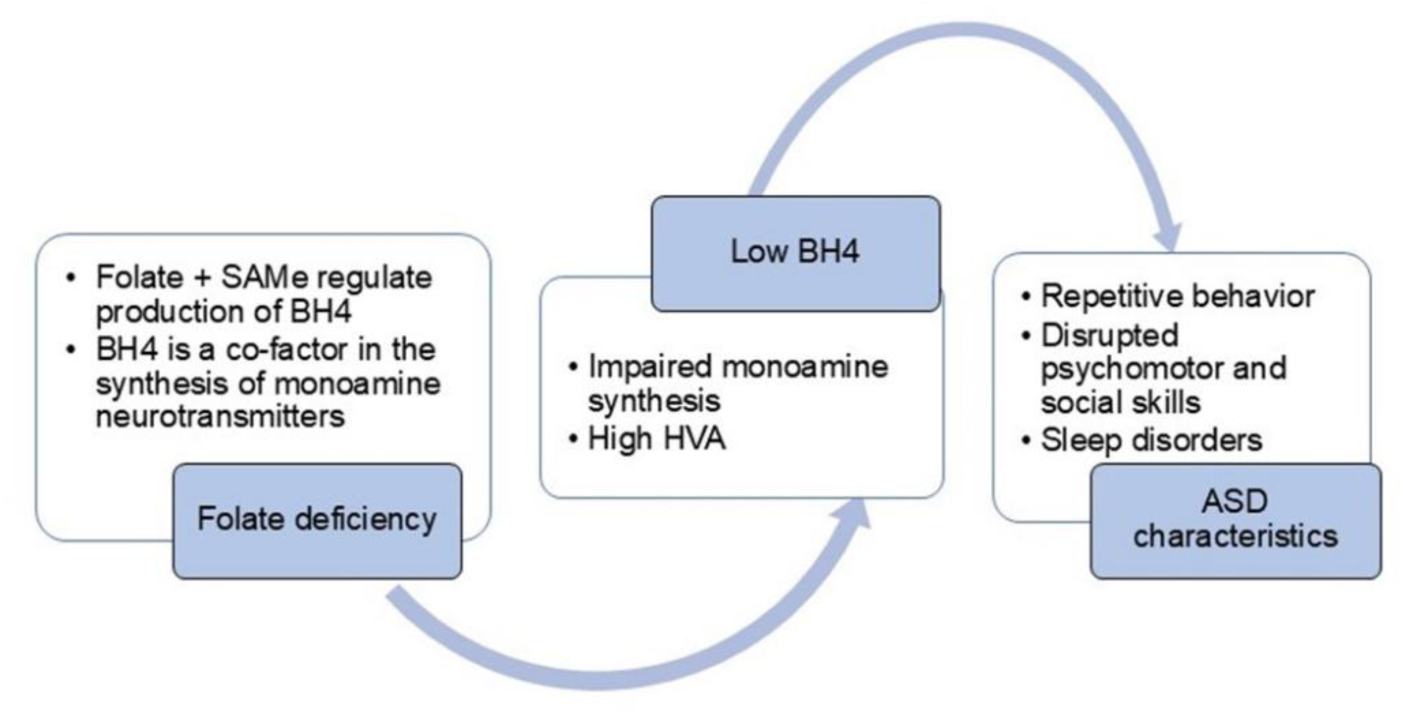
Folate deficiency linked to ASD characteristics.

#### 4.2.2 Anxiety and sleep disorder in ASD are linked to disturbed dopamine synthesis

Anxiety (Guerrera et al., 2022; van Steensel et al., 2011) and sleep disorders (Devnani & Hegde, 2015) are two of the most common ASD comorbidities. Research has shown that the dopaminergic system is involved in anxiety disorder (Dong et al., 2020) and in the regulation of the sleep cycle (Oishi & Lazarus, 2017). CSF studies establish that high HVA levels are found in ASD, indicating a disturbance in dopamine synthesis. Thus, anxiety and sleep disorders in ASD can be attributed to defects in the dopamine pathway.

#### 4.2.3 Tyrosine and tryptophan pathways are dysregulated in ASD

The tyrosine and tryptophan are the two important metabolic pathways (Figure 6) modulating mood, behavior, cognition, and neuroimmune interaction (Aquili, 2020). These pathways lead to the formation of metabolites like HVA, quinolinic acid, IAA, and 5-HIAA (Galla et al., 2021). Elevated HVA, low 5-HIAA, and quinolinic acid, as well as altered BH4 and NH_2_ levels, indicate dysfunction in tyrosine and tryptophan pathways.

**Figure 6:**
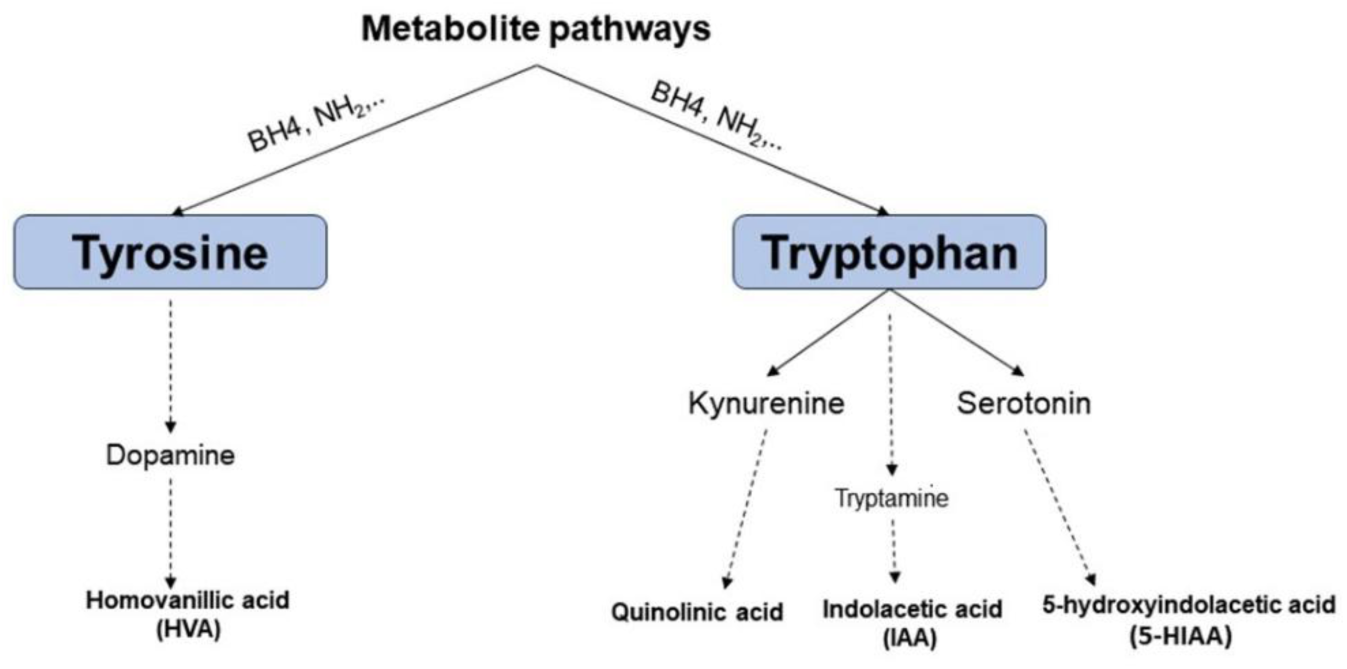
Tyrosine and Tryptophan pathways leading to the formation of HVA, Quinolinic acid, IAA, 5-HIAA.

### 4.3 Limitations of our review

This systematic review has some limitations. First, studies were systematically screened and reviewed by only one author (VS). However, the two authors regularly discussed the search and review procedures, the intermediate steps and outcomes of this process, and the problematic cases. The senior author (COR) selectively reviewed papers and areas with inconsistent or seemingly problematic results to ensure consensus between the two authors. Secondly, we did not consider co-morbidity in the ASD group while evaluating the study results. Finally, no review protocol was prepared in writing and registered before conducting the review.

### 4.4 Gaps in literature

We found several gaps in the literature related to sample size, age, and sex. In the studies presented, we found high variation in the sample sizes of the study and control groups. The study group sample sizes ranged from 3 to 10,000+ (Mohammad et al., 2024), and the size of the control group ranged from 0 (compared with previous studies) to 10,000+ (Mohammad et al., 2024). Table 7 shows that most of these studies have roughly between 10 and 29 participants per group. For a relationship to be detected with a 0.05 significance threshold at least 80% of the time with a sample size of N=29 per group, it needs to have an effect size of 0.75, which is rather large. Thus, most of these studies are not very sensitive and are expected to report non-significant results for small or medium effects.

**Table 7:** Number of studies for different sample sizes.

| Sample sizes | Study group | Control group |
| --- | --- | --- |
| Less than 10 | 12% (6/49) | 18% (9/49) |
| Between 10 and 29 | 47% (23/49) | 51% (25/49) |
| More than 50 | 24% (12/49) | 27% (13/49) |

We found multiple gaps regarding age in both the study and control groups. Multiple studies were conducted with a study group consisting of only children but a control group with both children and adults (Ahlsén et al., 1993; Rosengren et al., 1992; Than et al., 2023; Vargas et al., 2005). Similarly, in 9/49 (18%) of the ASD samples, there was a huge variation in the age range (5.3 – 19 years (Parker et al., 2018), 1 – 22 years (Nordin et al., 1998), 2 – 23 years (Perry, 1978), 3 – 42 years (Peterson et al., 2021b)), making it hard to do any reliable age-based analysis. Further, only 27% (13/49) of studies were carried out on ASD adults. Adult studies on ASD would help understand its progression and associated physiological changes with age. Furthermore, the research findings in ASD adults may motivate customized healthcare services to improve their quality of life. Also, many papers did not highlight the results by sex despite studying both males and females (Pardo et al., 2017; Ramaekers et al., 2020; Runge et al., 2020; Vargas et al., 2005). The reporting of research observations by biological sex is crucial because of significant sex differences in ASD (Napolitano et al., 2022). Consequently, sex differences in physiological traits should be systematically reported as they may indicate the need for different treatment and healthcare services. We also found that the proportion of females in the ASD group in each category (immune markers, folate, monoamines, proteins, EA CSF, and others) was between 16% and 32% only. Figure 7 shows the proportion of males and females in each category between 1977 and 2025.

**Figure 7:**
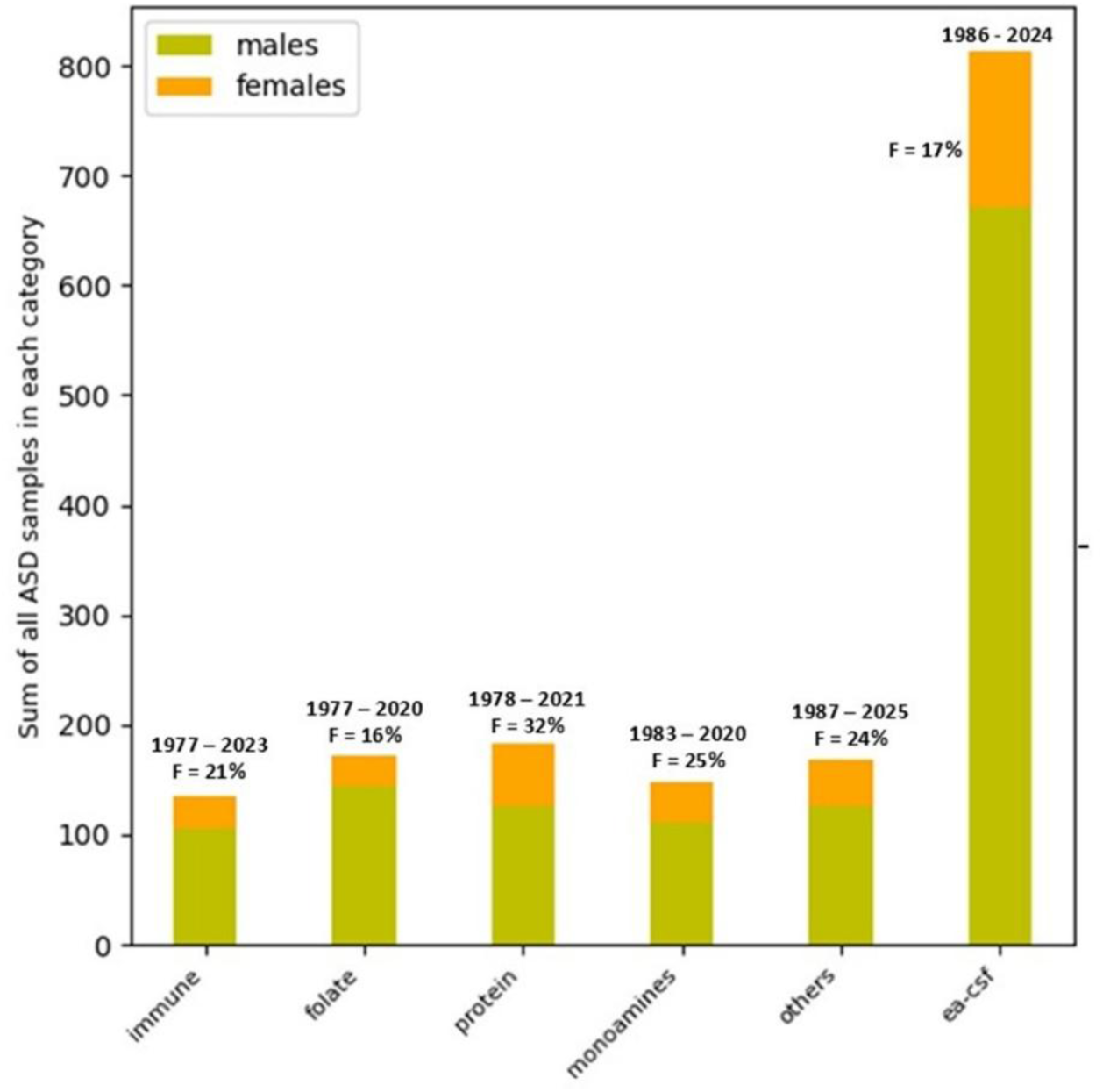
Proportion of females (F) in each category. The years of studies and the proportion of females for each category are mentioned at the top of the bar. Only the studies that reported samples by sex are included in the calculation.

## 5. Conclusions

In this systematic review, we found that CSF immune markers, proteins, axial-CSF, folate, oxytocin, and vasopressin change in ASD compared to the neurotypical population. These changes indicate neuroinflammation and an overactive innate immune system, dysregulated tyrosine, tryptophan, and dopamine pathways in ASD. We did not find significant changes related to immunoglobulin levels, beta-endorphins, nerve growth factor, indoleacetic acid, and serotonin.

We found many gaps in the research related to sample size, age, and sex. There was a high variability in the sample size of both the study and control group, with about 12% of the studies being conducted on very small samples (less than 10 participants per group). Also, most of the studies were conducted on children, making it difficult to establish the changes in CSF physiology in ASD due to age. We suggest more longitudinal studies be conducted to better understand the changes in CSF chemical properties in ASD by age. We observed that the ASD sample in 18% of the studies had a very wide age range, containing both toddlers and middle-aged adults (e.g., 3-42 years). In addition, studies were male-centric, with either all-male cohorts or very few female participants. Although the prevalence of diagnosed ASD has a 3:1 male-to-female ratio (Loomes et al., 2017), women are likely to be underdiagnosed (Loomes et al., 2017). This situation strongly motivates the need to study ASD in both sexes equally, to avoid adding to the injury of being underdiagnosed, the insult of being discriminated against in research and corresponding medical advances. We also found that even with participants of both sexes, the results were not reported by sex. In the future, studies should be conducted on a larger ASD cohort and include equal representation of females. Also, results should be reported for both males and females, or sex interaction effects should be reported.

With this systematic review, we found that the impairment of different interconnected systems characterizes ASD, leading to a significant change in CSF traits. This conclusion confirms our hypothesis that the properties and composition of CSF are altered in ASD. Since the number of studies is small compared to the wide range of CSF properties being targeted, more research needs to be done to establish CSF biomarkers for ASD.

## Data Availability

All data produced in the present work are contained in the manuscript.

## Author contributions

Vandana Srivastava: review and screening of papers, writing original draft, visualization, investigation. Christian O’Reilly: methodology, writing-review and editing, supervision.

## Acknowledgments

This work was supported by a pilot grant from the Carolina Autism & Neurodevelopment (CAN) Research Center (PI: Christian O’Reilly) and start-up funds (Christian O’Reilly) from the University of South Carolina. The funding bodies had no role in the study design, collection, analysis, or interpretation of data, the writing of the report, and the decision to submit the article for publication.

